# Cortical Hyperexcitability Shapes Large-Scale Brain Dynamics and Behavioral Outcome in Angelman Syndrome

**DOI:** 10.64898/2026.03.21.26348706

**Authors:** Gian Marco Duma, Margherita Bagnoli, Giulia Stefanelli, Camille Mazzara, Giovanni Pellegrino, Giovanni Mento, Pierpaolo Sorrentino, Lisa Toffoli, Fiorella Del Popolo Cristaldi, Lisa Antoniazzi, Rebecca Azzolini, Jacopo Dei Tos, Martina Baggio, Paolo Bonanni, Alberto Danieli

**Affiliations:** Scientific Institute IRCCS E.Medea, Epilepsy and Clinical Neurophysiology Unit, Conegliano (TV), Italy; NeuroDev Lab, Department of General Psychology, University of Padua, 35122 Padua, Italy; Institut de Neurosciences des Systémes, Aix-Marseille Université, 13005, Marseille, France; Department of Clinical Neurological Sciences, Schulich School of Medicine and Dentistry, Western University, London, ON, N6A 5C1, Canada; Università degli Studi di Napoli Parthenope, Dipartimento delle Scienze Mediche, Motorie e Del Benessere, Napoli, Italy

**Keywords:** Angelman syndrome, cortical excitability, functional connectivity, brain dynamics, sensory profile, high-density EEG

## Abstract

**Background:** Angelman syndrome (AS) is a rare neurodevelopmental disorder with characteristic electroencephalographic abnormalities caused by loss of function of the maternally inherited UBE3A gene. Converging evidence suggests a disrupted excitation–inhibition (E/I) balance towards hyperexcitability. However, noninvasive approaches capable of characterizing intrinsic cortical excitability and its relationship with large-scale brain dynamics in AS are still lacking. We used resting-state electroencephalography (EEG) to derive cortical excitability, testing the hypothesis that altered local E/I balance in AS is associated with instability of large-scale functional brain networks.

**Methods:** We recorded 7 minutes of task-free high-density EEG in 29 individuals with AS and 36 typically developing controls. Source-reconstructed cortical activity was used to compute the excitability index (EI), based on mean spatial phase synchronization in the gamma band. Dynamic functional connectivity was computed and summarized as fluidity index, which estimates temporal variability of network configurations. We assessed group differences and associations between EEG features, clinical variables and caregiver-reported questionnaires.

**Results:** AS participants showed increased EI in anterior cingulate, dorsolateral prefrontal, temporoparietal, and occipital regions. Fluidity was larger in AS across frequency bands, indicating greater network instability. EI positively predicted fluidity in widespread regions in AS, whereas the opposite pattern was observed in controls. Higher EI correlated with fewer antiseizure medications and with greater sensory-seeking behavior.

**Conclusions:** AS is characterized by cortical hyperexcitability coupled with unstable large-scale network dynamics. The EI provides a biologically meaningful marker linking intrinsic E/I imbalance to behavioral features and treatment-related variables

## Introduction

Angelman syndrome (AS) is a rare neurodevelopmental disorder caused by loss of function of the maternally inherited UBE3A gene, most commonly due to chromosomal deletions at 15q11–q13 (1). UBE3A encodes an E3 ubiquitin ligase that is selectively expressed from the maternal allele in neurons, regulating cellular protein homeostasis and influencing the expression of genes involved in neural development and functioning (1,2). Individuals with AS present with severe intellectual disability, speech impairment, motor disorder, epilepsy, sleep disturbances, and distinctive behavior characterized by a happy disposition, hyperactivity, short attention span, strong drive for social interaction and sensory-seeking behaviors (3,4). Electroencephalography (EEG) is characterized by abnormalities, particularly high-amplitude, rhythmic delta activity with posterior or anterior predominance, intermixed with epileptiform discharges, which are found in almost all individuals (5,6).

Converging preclinical evidence suggests that disruption of the excitation/inhibition (E/I) balance is a core mechanism underlying AS. In murine models, maternal loss of UBE3A is shown to alter multiple pathways involved in synaptic function and plasticity, including GABA transporter 1 (GAT1) degradation, activity of calcium- and voltage-dependent large-conductance potassium (BK) channels, and calcium/calmodulin-dependent protein kinase II (CAMK2) signaling (7–10). These effects vary across brain regions and neuronal populations, producing distinct neurophysiological and behavioral consequences (11). In particular, UBE3A loss in GABAergic neurons has been shown to induce AS-like EEG abnormalities and increased seizure susceptibility, whereas glutamatergic UBE3A loss primarily drives motor and behavioral abnormalities (12,13).

Additionally, 15q11–q13 deletions involve the loss of other genes contributing to AS pathophysiology, including the GABRB3–GABRA5–GABRG3 gene cluster encoding GABAA receptor subunits expressed in cortical and thalamocortical circuits. The resulting reduction in GABAergic neurotransmission has been proposed to underlie the more severe clinical presentation and some specific EEG features observed in deletion cases compared to other genetic etiologies (14). Together, these mechanisms are thought to shift the E/I balance toward cortical hyperexcitability, linking molecular deficits with the electrophysiological and clinical features of AS.

Traditionally, brain excitability in humans has been assessed using perturbational approaches such as transcranial magnetic stimulation (TMS), which primarily probe the motor cortex. Recent advances suggest that intrinsic brain activity may also carry signatures of underlying excitability (15,16). In particular, features derived from resting-state EEG have been proposed as noninvasive proxies of neuronal excitability, enabling broader spatial coverage and facilitating investigation in populations for whom stimulus-based paradigms are difficult, such as children and individuals with severe neurodevelopmental disorders (15–17).

Among resting-state markers linked to E/I balance, the aperiodic (1/f) exponent of the EEG power spectrum has received considerable attention, with steeper slopes typically interpreted as reflecting increased inhibition (18–21). However, recent studies have reported increased aperiodic exponents in AS, suggesting heightened inhibition, an observation that appears inconsistent with evidence of reduced GABAergic signaling in the disorder (22). This discrepancy raises important questions about the specificity and interpretation of current resting-state excitability markers in AS.

In the present study, we leveraged a metric derived from intrinsic neural dynamics, grounded in theoretical and empirical work linking spatial signal properties to E/I balance in epilepsy, to estimate cortical excitability in AS (16,23,24). We recorded resting-state brain activity using high-density EEG (128 channels) in individuals with AS and typically developing controls. Through source reconstruction, we estimated this excitability index at the regional level, enabling spatially resolved characterization of cortical E/I balance. We further examined how local excitability relates to large-scale network dynamics.

We hypothesized that this excitability index would provide a sensitive marker of AS-related neurophysiology and better align with known mechanisms of E/I balance disruption. Finally, we explored associations between regional excitability alterations and behavioral measures assessing sensory processing, aberrant behaviors, and social-communicative functioning, aiming to connect neurophysiological mechanisms and clinical variables

## Methods

### Participants

We enrolled patients with a diagnosis of Angelman syndrome (AS) who underwent high density EEG (hdEEG) between 2020 and 2025 at the Epilepsy and Clinical Neurophysiology Unit, IRCCS Eugenio Medea in Conegliano (Italy). Thirty-five cases were retrospectively screened and 5 patients were excluded due to poor data quality or missing behavioral questionnaires (N=1). The final sample consisted of 29 participants (age in years 12,82 ± 8,05 (mean ± SD),14 males, 15 females) with AS (25 with epilepsy; 24 deletion of the 15q11–q13 region; 5 non-deletional mutation. We also recorded data from a group of healthy controls (HCs) collected at the Department of General Psychology of Padova University. The HC group consisted of 36 subjects (age in years 10,33 ± 2,56 (mean ± SD); 16 F, 20 M) with no history of neurological and neuropsychiatric disorders. Patients’ demographic and clinical characteristics are provided in Table 1. The study protocol was conducted according to the Declaration of Helsinki and approved by the ethical committees (Comitato Etico Area Nord Veneto) and (Comitato etico della ricerca psicologica (area 17) - Dipartimenti/Sezione di Psicologia - Università di Padova, protocollo no. 387-a). All participants provided written informed consent to participate in the study.

**Table 1.** Sample demographic and clinical information. The continuous variables are reported as mean ± SD. ASMs, antiseizure medications.

| Patients with Angelman syndrome | Mean and Standard Deviation |
| --- | --- |
| Age (years) | $12.82 \pm 8.05$ |
| Age of onset (years) | $2.5 \pm 1.47$ |
| Duration of epilepsy (years) | $5.56 \pm 7.14$ |
| Lifetime number of ASMs | $3.04 \pm 1.88$ |
| Current number of ASMs at the EEG recording | $1.52 \pm 0.92$ |

### Resting-state EEG recording

Seven minutes of task-free hdEEG data was recorded with a R-Net EEG cap (128 sponge electrodes). Participants were sitting comfortably on a chair in a silent room, looking at the Inscapes video (25). We purposely used Inscapes video as it is validated for task-free neuroimaging studies and it represents a source of engagement, which is fundamental in a population with severe neurodevelopmental disorders. The signal was sampled at 500 Hz (Padova), 512 Hz (Conegliano), and referenced to the vertex.

### EEG preprocessing

Signal preprocessing was performed in EEGLAB 14.1.2 (26) according to a pipeline validated in previous works (27,28). The preprocessing pipeline included the following steps: (a) data resample to 250 Hz; (b) bandpass-filter (1 to 45 Hz) with a Hamming windowed sinc finite impulse response filter; (c) epoching (1 s-long epochs); (d) manual removal of epochs containing interictal epileptiform discharges (IEDs) and automatic removal of artifacted epochs using TBT EEGLAB plug-in; (e) removal of flat channels; (f) independent component analysis (40 independent components decomposition) (29), using the Infomax algorithm (30) as implemented in EEGLAB; (g) rejection of independent components related to eyes, muscle, channel noise and heart artifacts; (h) channel interpolation with spherical spline interpolation method (31); (i) re-reference to average; (il) re-concatenation of the epochs obtaining a continuous signal. For the patients group we rejected an average of 143.18 ± 58,82 (SD) (average of 34.04% of rejected epochs) epochs and an average of 23.52 ± 4.17(SD) (average of 58.8% of rejected components) components. For the control group we rejected an average of 2.43 ± 2.79(SD) (average of 0.59% of rejected epochs) and an average of 30.64 ± 4.80 (SD) (average of 76.6% of rejected components).

### Cortical source modelling

We used age-specific anatomy templates publicly available (32–34). The magnetic resonance images were segmented with Freesurfer (35) using CBRAIN (36) analytical cluster to obtain skin, skull and grey matter surfaces. Anatomical data were then imported in Brainstorm (37). The co-registration of the EEG electrodes with brain MRI was performed using Brainstorm, considering anatomical landmarks. Manual co-registration corrections were applied as needed. The cortical mesh was downsampled at 15002 vertices and the three boundary element model (BEM) surfaces were reconstructed (inner skull, outer skull and head). We computed the BEM OpenMEEG plugin (38,39) implemented in Brainstorm. We used the weighted minimum norm as an inverse solution method, with Brainstorm’s default parameter settings (40). Vertex-wise activity was then downsampled to the Desikan-Killiany atlas (41).

### Excitability Index Computation

The excitability index, originally proposed and validated in epileptic populations, quantifies mean spatial phase synchronization and, particularly within high-gamma frequency bands (23,24). It showed a strong correlation with cortical excitability as assessed using perturbational methods and as modulated by antiseizure medications in patients with epilepsy (16,23,24). This excitation–inhibition (E/I) measure is conceptually related to the widely used phase-locking value (PLV), with the key distinction that it is computed across spatial dimensions rather than over time. Following prior recommendations, we focused on the gamma band (30-45 Hz).

### Functional Connectivity

Dynamic functional connectivity (FC) was estimated at the source level using lagged coherence (lagCoh). Unlike magnitude-squared coherence, lagCoh removes the instantaneous (zero-phase) component, thereby reducing the effects of volume conduction and common referencing and providing a more robust estimate of genuine time-delayed interactions (42–44). Connectivity was computed using Brainstorm functions.

Connectivity was calculated using a sliding-window approach (8 s window, 90% overlap). FC was estimated in five canonical frequency bands: delta (2–4 Hz), theta (4–7 Hz), alpha (8–12 Hz), beta (13–30 Hz), and gamma (30–45 Hz), yielding a connectivity tensor of size N (brain regions) × N × T (number of windows) × F (frequencies).

### Fluidity

Dynamic functional connectivity matrix (dFC) was obtained by correlating the upper triangular elements of lagCoh matrices from consecutive windows, capturing the temporal evolution of inter-regional synchronization:

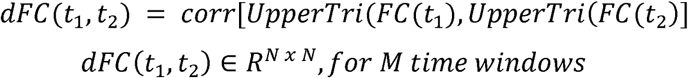

Fluidity was defined as the variance of the upper triangular values of the dFC matrix after removing overlapping windows

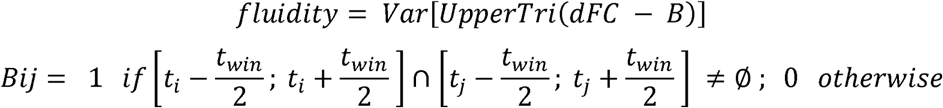

with the diagonal offset to compute fluidity given by the following formula

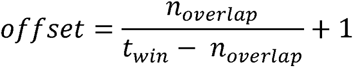

This metric reflects the temporal variability of functional connectivity and was interpreted as an index of the brain’s dynamic adaptability (45,46). The analytical pipeline is represented in Fig.1.

**Figure 1.**
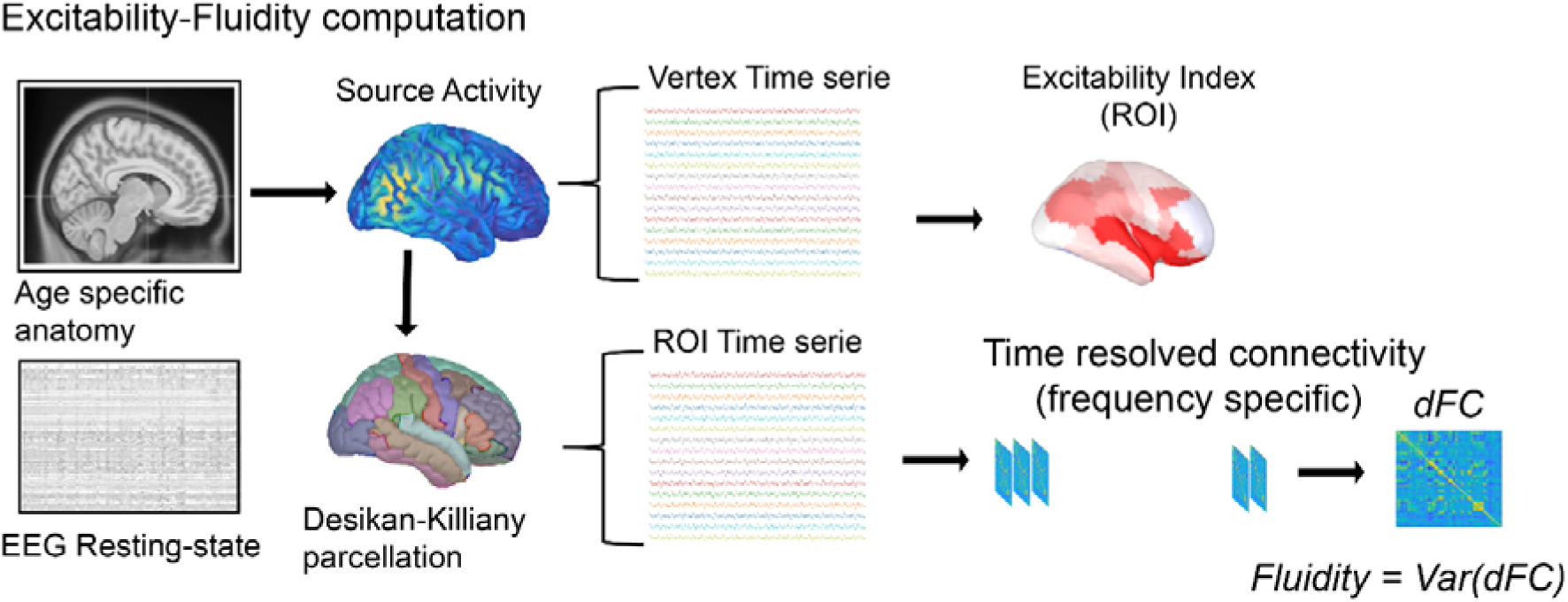
Analyses pipeline for excitability and fluidity. The figure shows the analysis pipeline from source reconstructed resting state electroencephalography (EEG) to obtain local measure of cortical excitability (Excitability Index) and global measure of network dynamics (Fluidity).

### Statistics

Regional differences in excitability values were evaluated using a permutation t-test approach (5000 permutations) with false discovery rate (FDR) correction for multiple comparison across the number of regions. Differences in fluidity were assessed using a generalized linear model (GLM) in R, using gamma family distribution, which best approximated fluidity distribution with the following model: Fluidity ∼ Group*Frequency_band. Relationship between fluidity and excitability index was investigated iteratively for each brain parcel with a mass-univariate GLM separately for each frequency: Fluidity ∼ Excitability*Group. P-values of the β of the model for each factor and the interaction effect were corrected with FDR. Correlations with questionnaires and clinical variables were computed using Spearman’s correlation, with FDR correction.

### Caregiver-reported questionnaires

#### Aberrant Behavior Checklist (ABC)

The Aberrant Behavior Checklist (ABC) (47) is a 58-item caregiver-rated measure of maladaptive behavior in neurodevelopmental disorders, with five subscales: Irritability, Social Withdrawal, Stereotypy, Hyperactivity, and Inappropriate Speech.

#### Social Communication Questionnaire (SCQ)

The Social Communication Questionnaire (SCQ) (48) is a 40-item caregiver-report screening tool for autism-related social communication difficulties and restricted/repetitive behaviors, derived from the ADI-R.

#### Sensory Profile 2 (SP-2)

The Child Sensory Profile 2 (SP-2) (49) is a 86-item caregiver questionnaire assessing sensory processing across daily contexts. It provides four sensory pattern scores (Seeking, Avoiding, Sensitivity, Registration) and sensory system subscales including Auditory, Visual, Touch, Movement, Body Position, and Oral processing, as well as Behavioral, Emotional–Social, and Attention domains. The Table 2 shows the descriptive statistic of the questionnaire scores.

**Table 2.**
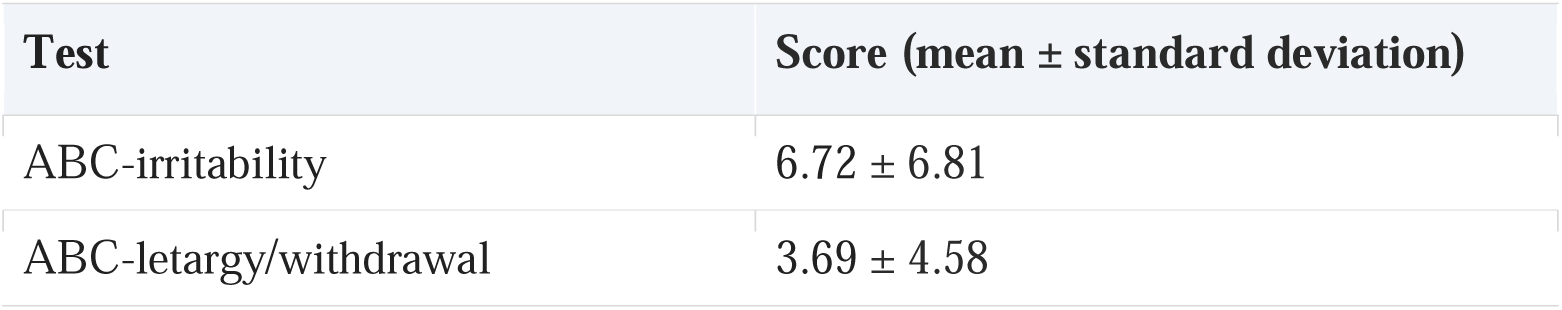
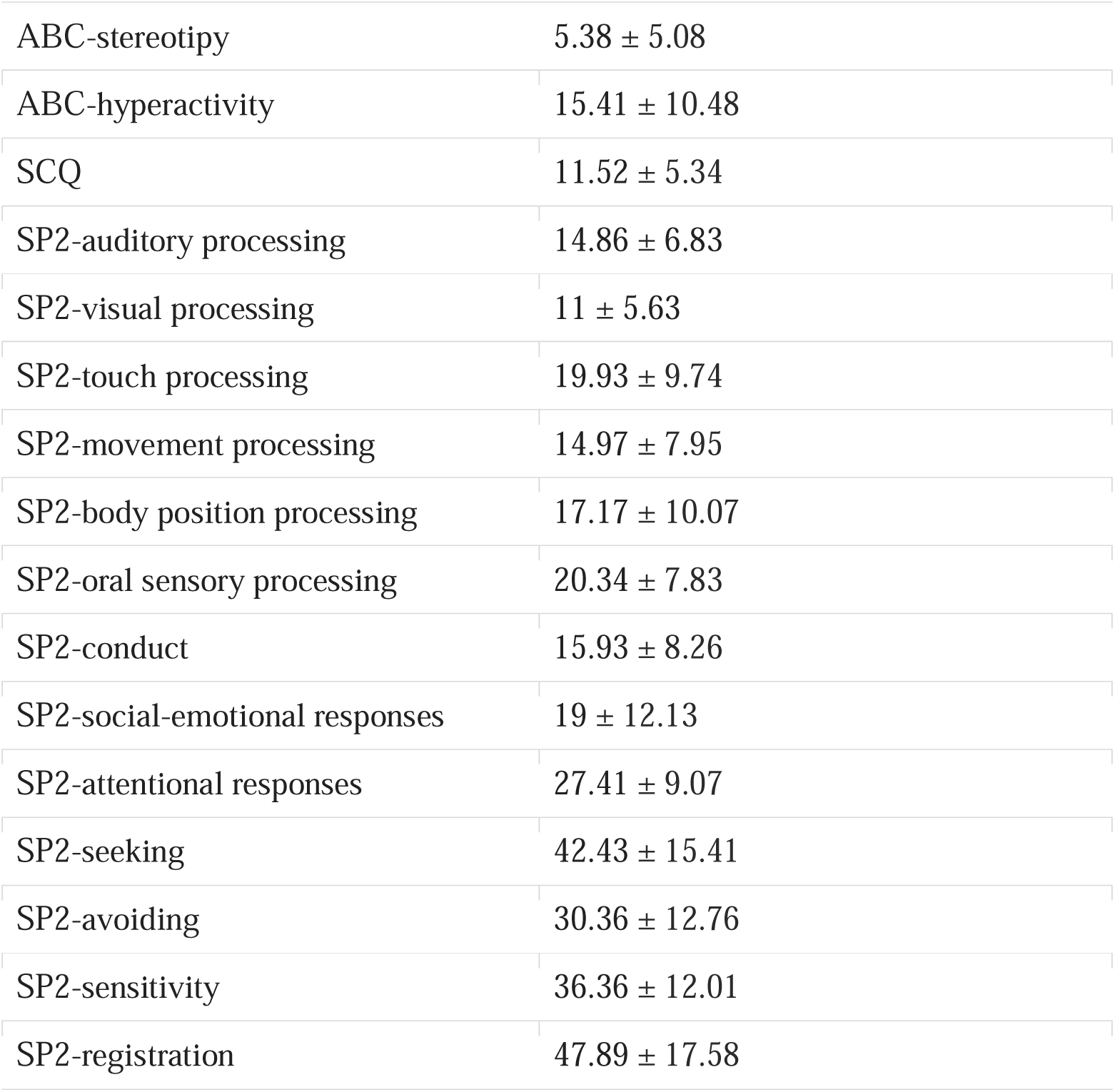
Descriptive statistics for all subscales of the parent-reported questionnaires. Continuous variables are presented as mean ± standard deviation (SD). Abbreviations: ABC, Aberrant Behavior Checklist; SCQ, Social Communication Questionnaire; SP-2, Sensory Profile 2.

## Results

### Alterations in local excitability and network configuration

We identified increased values of the excitability index (EI) in patients with AS as compared to the control group. This effect was expressed bilaterally including fundamental hubs of the default mode network such as anterior cingulate, prefrontal dorsolateral cortices and cuneus (*p_FDR_ < .05*). Additionally, EI increases were also detected in temporoparietal junction and tempol giri (see Fig.2A). When looking at the fluidity index the GLM analysis evidenced a significant effect of the Group factor (*F* = 7.710; *p* = .005), but no effect on the frequency band (*F* = .082; *p* = .987) and the interaction Group x Frequency (*F* = .368; *p* = .937).

**Figure 2.**
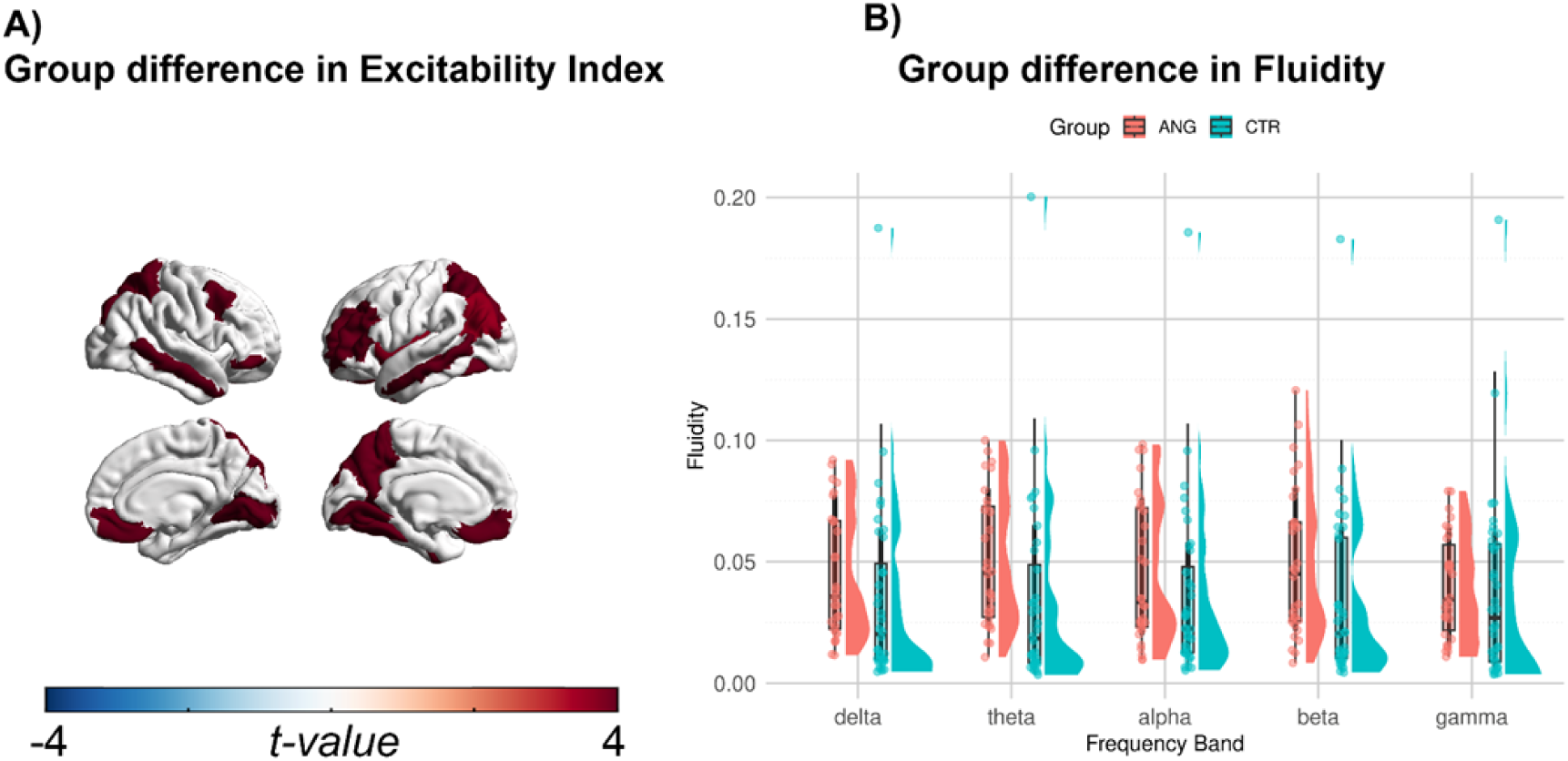
Group difference in Excitability index and fluidity. Panel A shows the spatial differences in the comparison of the excitability index between patients with Angelman syndrome vs. Controls. Panel B shows the group differences in fluidity across the five canonical frequency bands between Angelman patients (pale red) and controls (teal).

When investigating the relationship between local EI values and the global organization of the network dynamics (fluidity) with a mass-univariate GLM approach we observed a significant interaction between fluidity and excitability. Specifically, after FDR correction we identified positive significant β (*p_FDR_* < .05) engaging a wide range of brain regions across all the frequency bands excluding gamma. In the Fig.3A we report the results at delta, which is the brain rhythm which alterations characterize AS, and alpha which represent the dominant rhythm of physiological brain activity during a task-free condition. Results in other frequencies are reported in the supplementary material (see Supplementary Fig1A). By contrast, we observed a negative relationship between EI and fluidity at the level of the parahippocampal cortex.

**Figure 3.**
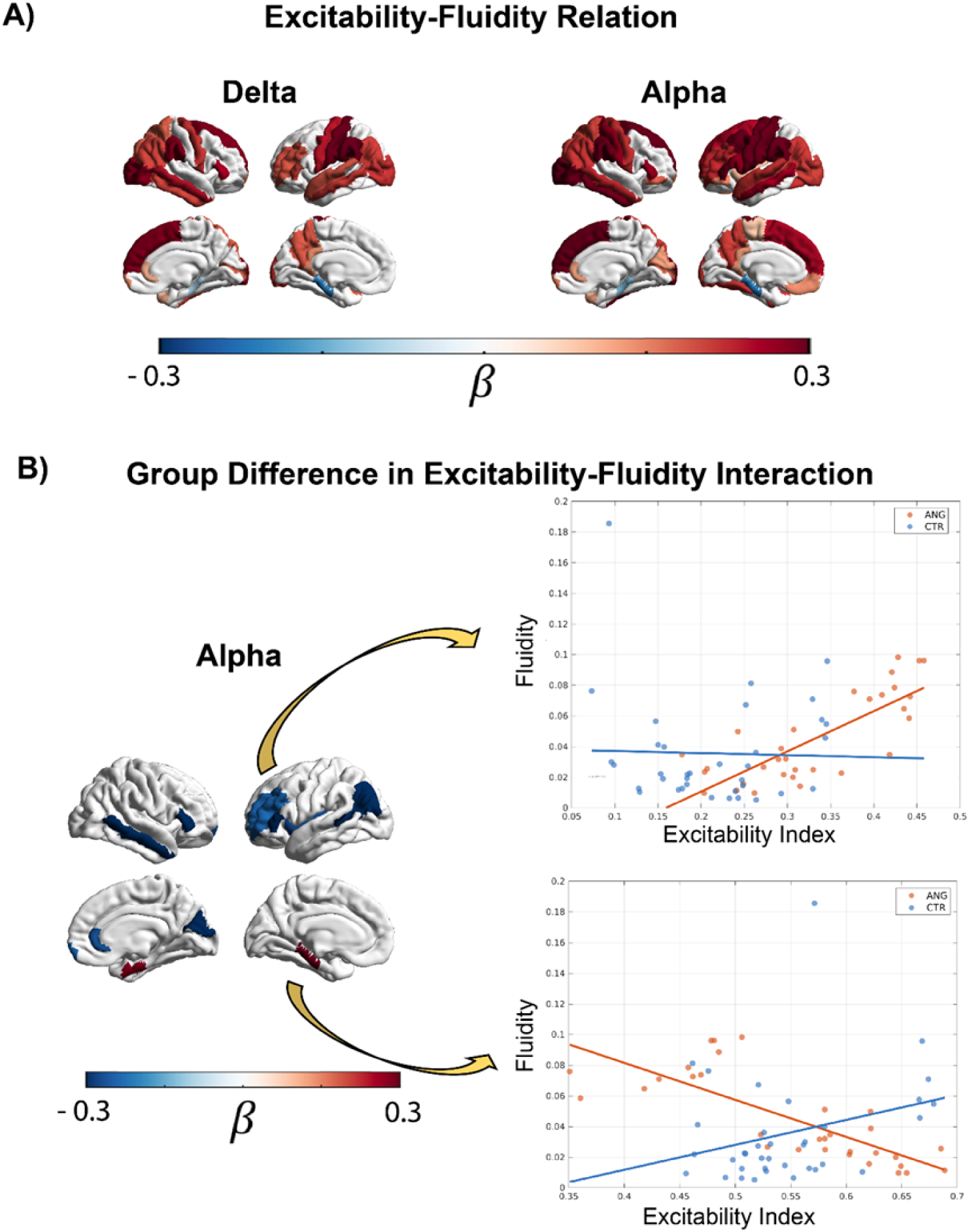
Mass-Univariate generalized linear model effects. A) Spatial distribution of the main effect of the excitability index in the generalized linear model (GLM). The colored areas represent the brain regions with significant beta values after false discovery rate (FDR) correction. This panel shows the effect only in the delta and alpha which represent the oscillatory rhythms with specific alterations in Angelman patients, and the dominant rhythm of the brain during resting condition, respectively. B) *Interaction effect of the GLM*. The colored brain areas represent the regions with significant interaction effects. The blue regions, (negative beta) indicate that the relationship between excitability and fluidity is more positive in the Angelman group compared to controls. On the right, the scatter plot and the regression lines show the interaction effect from the left rostral middle frontal area of the Desikan-Killiany atlas (scatter plot: orange = Angelman Syndrome; blue = healthy controls). The red regions (positive beta values) display an opposite direction of the effect, the excitability index-fludity relationship more positive in the control group. The scatter plot and regression lines in the right lower corner display the interaction effect in the left parahippocampal cortex.

When characterizing the difference between groups by looking at the Excitability*Group interaction we observed negative significant β (*p_FDR_* < .05) at the level of dorsolateral prefrontal and anterior cingulate cortices, precuneus, and temporoparietal junction (see Fig.3B). When looking at the slopes, we observed that the two groups are characterized by an opposite effect. In patients with AS larger EI is associated with larger fluidity, while the opposite pattern is present in the controls (see Fig.3B). These effects are present across multiple frequencies. In the Fig.3B we report results in alpha bands. The remaining frequencies are reported in supplementary materials (see Supplementary Fig1.B). Finally, the parahippocampal cortex showed an opposite effect, with larger fluidity associated to lower EI values in AS (see Fig.3B)

### Correlations between EI and clinical variables

To investigate the clinical relevance of the excitability index (EI), we examined its association with behavioral and sensory phenotypes assessed through caregiver-reported questionnaires. Based on the known sensory-seeking profile of individuals with Angelman syndrome, we specifically hypothesized that increased cortical excitability would be associated with altered sensory processing. Consistent with this hypothesis, we observed a significant positive correlation between EI values and sensory seeking scores of the Sensory Profile 2. This relationship was localized to the right anterior cingulate and dorsolateral prefrontal cortices (*p_FDR_*< .05), indicating that higher cortical excitability in these regions was associated with increased sensory-seeking behavior in patients with AS (Fig. 4C). In addition, EI values were also related to clinical epilepsy variables. Specifically, we found a negative correlation between EI and the number of antiseizure medications (ASMs) taken both across the lifespan and at the time of EEG recording (Fig. 4A–B), such that patients treated with a higher number of ASMs exhibited lower EI values.

**Figure4.**
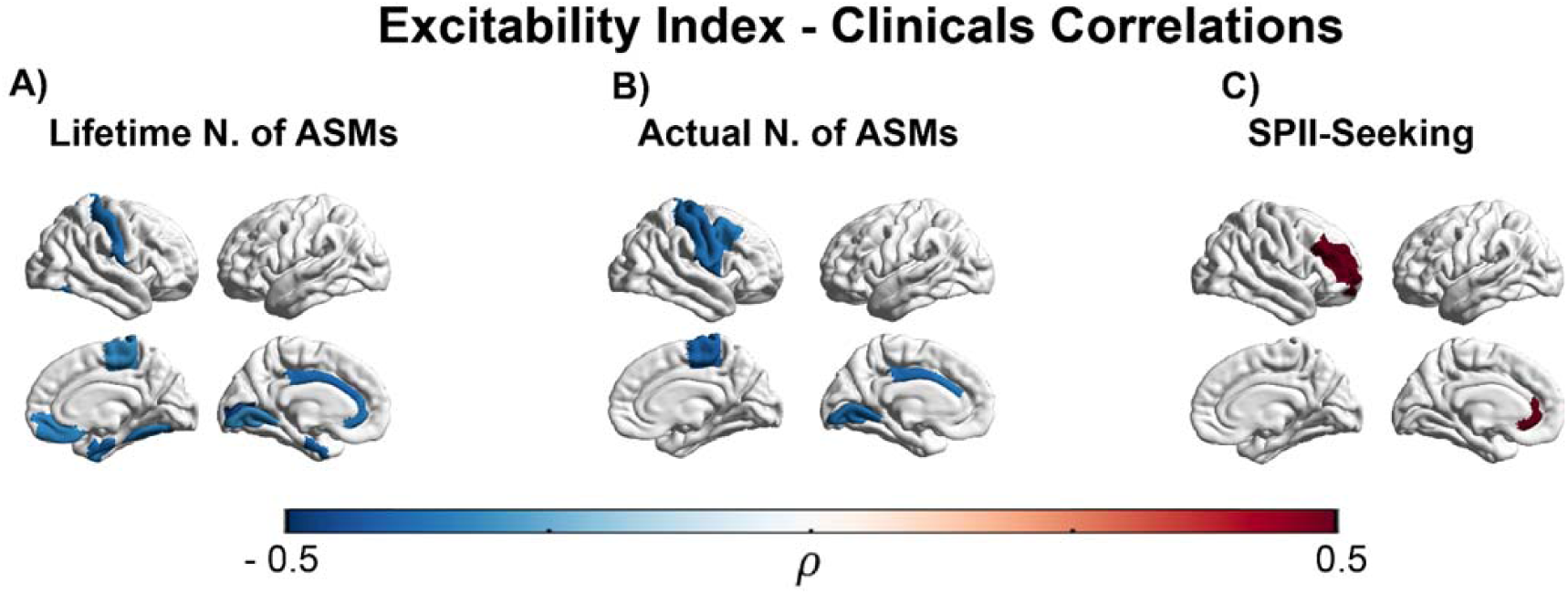
Correlations between excitability index, clinical and behavioral variables. Panels A and B show the correlation between excitability index and the total number of antiseizure medications (ASMs) taken during the lifetime and at the moment of the EEG recording. Panel C shows the correlation with the seeking scale of the Sensory Profile II. Colored regions indicate the Spearman’s of the significant brain areas after FDR correction

## Discussion

The present study provides novel evidence that Angelman syndrome (AS) is characterized by widespread alterations in intrinsic cortical excitability and its coupling with large-scale brain dynamics. By combining a spatially resolved excitability index with measures of dynamic functional connectivity, we were able to link local excitation–inhibition balance with global network organization and clinically relevant behavioral features.

First, we observed a robust bilateral increase in the excitability index (EI) in individuals with AS compared to controls, involving core regions of the default mode network, including the anterior cingulate cortex, dorsolateral prefrontal cortex, cuneus, temporoparietal junction, and temporal gyri. This pattern is consistent with the known pathophysiology of AS, in which synaptic dysfunction and reduced GABAergic signaling are thought to bias neural circuits toward hyperexcitability (7–9,11–13,50). Notably, EI increases were prominent in higher-order associative cortices, suggesting that excitation–inhibition imbalance in AS operates at a systems level and affects integrative networks. These regions are key hubs for internally oriented cognition, multisensory integration, and cognitive control (51–53). Altered excitability within this distributed network may therefore influence processes related to sensory integration, behavioral regulation, and information coordination, with potential widespread effects on large-scale brain functioning (54).

At the global level, individuals with AS showed consistently higher brain fluidity across frequency bands, indicating a more variable and less stable configuration of large-scale network dynamics. Rather than reflecting enhanced flexibility per se, this pattern may indicate a system that more frequently shifts between transient states, potentially as a consequence of altered excitation–inhibition balance. Increased excitability may render neural activity less constrained, promoting more frequent reconfiguration of functional connectivity. Such instability is in line with the clinical phenotype of AS, characterized by fluctuations in attention, behavior, and sensory responsivity (55). Importantly, our results revealed a clear relationship between local excitability and global fluidity: across widespread cortical regions, higher EI values were associated with greater network variability, suggesting that local excitation–inhibition properties play a key role in shaping the stability of large-scale brain organization (56,57).

A key finding emerged from the Excitability × Group interaction. In associative hubs, including the dorsolateral prefrontal cortex, anterior cingulate cortex, precuneus, and temporoparietal junction (58), we observed opposite coupling patterns in AS and controls. In AS, higher excitability was associated with increased fluidity, whereas in controls it tended to relate to more stable network configurations. These regions support cognitive control, salience processing, multisensory integration, and internally oriented cognition and act as convergence points for long-range communication (58,59). The divergent coupling suggests that, in the typical brain, local excitability within these hubs may contribute to stabilizing coordinated activity (60,61), whereas in AS—where baseline excitability is already elevated—further increases may push the system toward a more unstable regime, contributing to widespread variability in network dynamics

An additional noteworthy finding concerns the inverse relationship observed between excitability and fluidity in the parahippocampal cortex, where higher EI was associated with lower network variability in AS. In the typical brain, this region may facilitate transitions between large-scale network states through its connections with hippocampal and retrosplenial structures and its role in coordinating slow oscillatory rhythms involved in memory-related processing and large-scale temporal organization of neural activity (62,63). In AS, where the broader cortical system already operates in a hyperexcitable and dynamically unstable regime, the functional role of the parahippocampal cortex may shift. The reversal of the EI–fluidity relationship in this region may therefore reflect a compensatory or stabilizing mechanism within medial temporal structures. However, this interpretation should be confirmed by future studies specifically targeting structural and functional alterations of the medial temporal lobe in AS.

The clinical relevance of the EI metric is further supported by its relationship with pharmacological and behavioral variables. We found a negative correlation between EI and the number of antiseizure medications (ASMs), both lifetime and current. This finding is consistent with the pharmacodynamic effects of ASMs, many of which enhance inhibitory neurotransmission or reduce neuronal excitability (64). The association suggests that EI may be sensitive to treatment-related modulation of cortical excitability, reinforcing its validity as a physiologically meaningful index.

In addition, we observed a positive correlation between EI values in the right dorsolateral prefrontal and anterior cingulate cortices and the seeking behavior dimension of the Sensory Profile 2. Individuals with higher excitability in these regions tended to display stronger sensory-seeking tendencies. These areas are involved in motivational regulation, salience attribution, and goal-directed exploration (65,66), and increased excitability within these circuits may heighten responsiveness to sensory input or drive increased engagement with environmental stimuli. In AS, sensory-seeking behaviors may represent an adaptive strategy for interacting with the environment in the context of severe communication and cognitive limitations, providing alternative channels for regulating arousal and expressing needs.

Our results should be interpreted with caution due to the relatively small sample size which may limit statistical power and generalizability. Replication in larger and multicenter cohorts will therefore be important to confirm the robustness of the observed excitability–network relationships and to further explore their clinical implications.

At the same time, an important strength of the present study lies in the feasibility of acquiring high-density EEG recordings in this population. The use of sponge-based electrode systems allowed rapid and well-tolerated cap application, minimizing discomfort and preparation time. Moreover, the combination of high-density EEG with the Inscapes naturalistic video (25) paradigm proved highly manageable for individuals with AS, who were able to tolerate the recording session and remain engaged with the visual stimulus. This setup represents a particularly promising approach for neurophysiological investigations in populations with severe neurodevelopmental disorders, where compliance with traditional experimental paradigms is often challenging. Beyond research applications, this methodology may therefore offer valuable opportunities for longitudinal monitoring, biomarker development, and the evaluation of treatment effects in clinical settings.

Taken together, these findings highlight the translational potential of excitability and network dynamics metrics derived from resting-state EEG. The sensitivity of EI to pharmacological modulation and its association with behavioral features suggest that it may represent a promising biomarker for tracking disease mechanisms and monitoring therapeutic interventions. More broadly, the coupling between local excitability and global network dynamics provides insight into a key dimension of brain organization altered in Angelman syndrome, linking physiological alterations with large-scale brain functioning and behavior. Longitudinal studies will be important to determine whether these measures can inform clinical trajectories and support future therapeutic trials.

## Supporting information

see Supplementary Fig.1A/B

## Data Availability

Data will be availaThe data that support the findings of this study are available on request to the corresponding author. The raw data are not publicly available due to privacy or ethical restrictionsble

## Acknowledgments and Disclosures

The study was funded by Ricerca Corrente 2026 from the Italian Ministry of Health; NSERC Discovery Grant, NSERC RTI Grant, AMOSO Opportunity Fund, Western CNS Internal Competition Grant, Lawson Internal Competition Grant, Western Seed Fund for CIHR Success, Western Start-up Grant. The funders had no role in study design, data collection, data analysis, data interpretation, writing, approval or submission of this manuscript.

GMD and MB had full access to all of the data in the study and took responsibility for the integrity of the data and the accuracy of the data analyses. GMD, MB, and AD were responsible for study concept and design.

LA, GM, GS, LT, FDPC, RA, JDT, MB were responsible for data acquisition. GMD, MB, GP, PB and AD were responsible for drafting the manuscript. GMD, GM, AD, GP, PS, CM and PB were responsible for making critical revisions to the manuscript for important intellectual content. GMD, MB and CM were responsible for statistical analysis. GM, PS, GS, PB and AD provided administrative, technical, or material support. PS, GP, PB and AD were responsible for study supervision. All the authors report no biomedical financial interests or potential conflicts of interest. The data that support the findings of this study are available on request to the corresponding author. The raw data are not publicly available due to privacy or ethical restrictions.

