## Supplementary material for "Cortical Hyperexcitability Shapes Large-Scale Brain Dynamics and Behavioral Outcome in Angelman Syndrome": see Supplementary Fig.1A/B

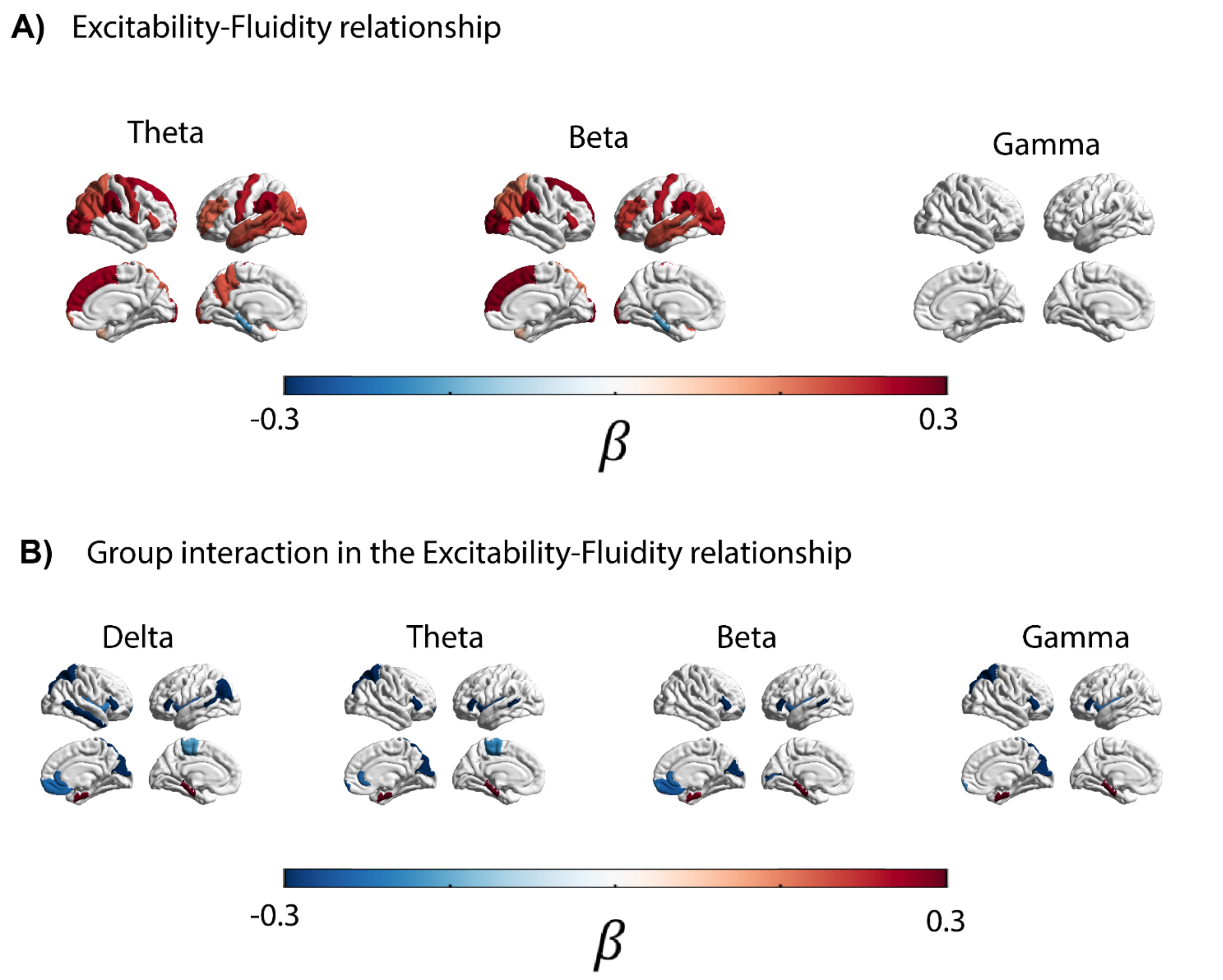


**Supplementary Figure 1. Mass-Univariate generalized linear model effects.** A) Spatial distribution of the main effect of the excitability index in the generalized linear model (GLM). The colored areas represent the brain regions with significant beta values after false discovery rate (FDR) correction. This panel shows the effect only in theta, beta and gamma bands. B) *Interaction effect of the GLM*. The colored brain areas represent the regions with significant interaction effects. The blue regions, (negative beta) indicate that the relationship between excitability and fluidity is more positive in the Angelman group compared to controls. The red regions (positive beta values) display an opposite direction of the effect, the excitability index-fluidity relationship more positive in the control group. The panel shows the interaction effect in delta, theta, beta and gamma band, emphasizing consistency fo regions across frequency bands.
